# Applying an Intersectional Approach to Examine the Associations between Sexual Minority Stressors, Resilience, and Substance Misuse Disparities among a National Sample of Sexual Minority Populations

**DOI:** 10.1101/2025.01.21.25320897

**Authors:** Chen Zhang, Shan Qiao, Wonkyung Chang, Yu Liu

## Abstract

**Objectives:** Our study aims to investigate the associations between sexual minority stressors, resilience factors, and substance misuse outcomes, using an intersectional framework to examine heterogeneities across sexual minority populations (SMPs). The study hypothesized that resilience factors would mitigate the adverse effects of minority stressors on alcohol and drug misuse risks.

**Design:** The current study employed a secondary data analysis strategy to analyze a cross-sectional data using Bayesian hierarchical modeling and Multilevel Analysis of Individual Heterogeneity and Discriminatory Accuracy (MAIHDA).

**Setting:** Data were obtained from the Generation Study (Wave 1), a nationally representative cohort of SMPs in the United States.

**Participants:** The study included 1,518 participants aged 18–59. Combinations of education, birth sex, sexual orientation, income, and race/ethnicity defined intersectional strata. Participants were selected based on self-reported demographic and behavioral data.

**Primary and Secondary Outcome Measures:** Primary outcomes were the risks of alcohol use disorder (AUD) and drug use disorder (DUD), measured using validated scales (AUDIT and DUDIT). Explanatory variables included resilience factors (social support, life satisfaction, social well-being, community connectedness) and sexual minority stressors (everyday discrimination, perceived stigma, healthcare stereotype threat, internalized homophobia).

**Results:** Bayesian modeling identified everyday discrimination as the strongest predictor of AUD (Estimate = 0.10, 95% CI [0.01, 0.18]) and DUD (Estimate = 0.23, 95% CI [0.15, 0.32]). Perceived stigma was negatively associated with AUD risk (Estimate = –0.06, 95% CI [-0.12, 0.00]), and social well-being was inversely associated with DUD risk (Estimate = –0.17, 95% CI [-0.23, –0.10]). MAIHDA revealed that intersectional strata contributed to 1.61% of the variance in AUDIT scores and 1.88% in DUDIT scores. Including resilience and stressors in the model explained 93.6% of the strata-level variance for AUDIT and 34.45% for DUDIT, underscoring the significant impact of these factors on substance misuse disparities.

**Conclusions:** The findings highlight the complex interplay between resilience, stressors, and intersectional identities in substance use behaviors among SMPs. Using an intersectional framework and MAIHDA can enhance understanding of heterogeneities in substance misuse across strata and inform targeted, equity-focused interventions to reduce disparities. Further research should explore longitudinal effects and intervention development.

## INTRODUCTION

Substance use disorders, including alcohol and drug misuse, remain critical public health challenges globally, with disproportionate burdens observed among sexual minority populations (SMPs).^1,2^ Evidence consistently demonstrates higher prevalence rates of alcohol and drug use among SMPs compared to their heterosexual counterparts, likely driven by complex interplays of social stigma, discrimination, and unique stressors experienced across diverse, intersectional identities.^3^

The elevated prevalence of substance use among SMPs has been linked to minority stress theory, which explains how stigmatization and discrimination create chronic stressors and increase health risks.^4^ Specifically, SMPs frequently experience unique stressors, including internalized stigma, healthcare stereotype threats, and everyday discrimination.^4,5^ These stressors can increase susceptibility to substance use and limit access to protective factors (e.g., social support and community connectedness). These stressors further exacerbate the use of substances as a coping mechanism, resulting in serious health issues such as increased sexual risk through condomless sex, self-harm, depression, and poor quality of life.^6^ Conversely, resilience factors like social well-being and self-satisfaction can potentially mitigate the adverse effects of these stressors.^7,8^ However, existing studies often examine these factors in isolation, failing to account for the intersectionality of social identities that shape these experiences.^4,6,9^ The intersectionality framework moves beyond focusing on a single identity, helping us understand the complex experiences of individuals shaped by multiple identities (e.g., race, sexual orientation).^9–11^ Incorporating intersectionality into health research can provide a more nuanced understanding of disparities and address the unique needs of diverse subgroups within SMPs.^12^

In addition to the lack of studies exploring how intersectional identities and social status affect substance abuse experiences, current research on alcohol and drug use behaviors among SMPs faces several methodological limitations. (a) limited sample size as the result of the multilayered stratification process; (b) compromised model parsimony and scalability due to the high dimensionality of strata intricacies; (c) interpretability challenges due to the complex analytical procedures of synergistic or antagonistic interactions of identified strata, and (d) the strata-level averages may obscure individual’s risk profiles due to the homogenization of diverse experiences within groups, masking the significant variability and nuances in health determinants and outcomes among individuals.^13–16^ These issues obscure critical distinctions in health determinants among diverse individuals, impeding a comprehensive understanding of health disparities. Therefore, we need an advanced statistical model to examine the varied experiences within and between intersecting socioeconomic strata.

To overcome identified methodological challenges,^13–16^ we advocate for applying the Multilevel Analysis of Individual Heterogeneity and Discriminatory Accuracy (MAIHDA) to examine group averages alongside variations at multiple levels, both within and across groups.^17,18^ MAIHDA has emerged as a powerful tool for advancing intersectionality theory^15,18^ in quantitative research, allowing researchers to quantify the discriminatory accuracy of social strata in predicting health outcomes. By incorporating random intercepts and slopes, this approach captures the variability of stressor and resilience effects across intersectional groups, providing a more nuanced understanding of health disparities.^18–20^ In the context of SMP research, MAIHDA can uncover how combinations of social identities shape alcohol and drug use behaviors, revealing patterns that traditional models fail to capture.^15,18^

The primary objective of this study is to examine how intersectionality impacts the associations between stressor and resilience profiles and substance use behaviors (i.e., alcohol and drug use) among SMPs. Using data from a nationally representative sample of the Generation Study,^21^ this research addresses the following questions: (1) How do resilience factors (e.g., social support, self-satisfaction, social well-being, community connectedness) and stressors (e.g., everyday discrimination, internalized homophobia, healthcare stereotype threat) relate to alcohol and drug use outcomes in SMPs? (2) To what extent do intersectional strata, defined by education level, sex, sexual orientation, income, and race/ethnicity, influence these relationships? (3) How do between-strata heterogeneity and within-strata variability contribute to understanding substance use behaviors in SMPs? By integrating intersectionality theory and MAIHDA, this study aims to fill critical gaps in the literature, advancing our understanding of the structural and social mechanisms driving substance use disparities among SMPs.

## METHODS

### Study Design and Data Collection

This study employed a secondary data analysis of cross-sectional data from the Generation Study, a nationally representative cohort investigating stress, health outcomes, and substance use among sexual minority populations (SMPs) in the United States.^21^ The Generation Study recruited participants aged 18-59 from three age cohorts using a dual-frame random sampling procedure.^21^ The final analytic sample consisted of 1,518 participants, with sociodemographic diversity in gender, sexual orientation, race/ethnicity, income, and education levels. Data were collected through self-administered surveys, capturing detailed measures of substance use, sexual minority stressors, and resilience factors.

### Key Variables

The primary outcomes of interest were alcohol use, assessed with the Alcohol Use Disorders Identification Test^22–24^ (AUDIT [Cronbach’s alpha=0.790]), and drug use, measured using the Drug Use Disorders Identification Test^25,26^ (DUDIT [Cronbach’s alpha=0.856]). Key explanatory variables included *resilience factors* (e.g., social support^27^ [Cronbach’s alpha=0.925], life satisfaction^28^ [Cronbach’s alpha=0.905], social well-being^29^ [Cronbach’s alpha=n/a], and community connectedness^30^ [Cronbach’s alpha=0.863]), *stressors* (e.g., everyday discrimination^31^[Cronbach’s alpha=0.909], perceived stigma^32,33^ (Cronbach’s alpha=0.902), healthcare stereotype threat^34^ [Cronbach’s alpha=0.902], and internalized stigma^35^ [Cronbach’s alpha=0.902]). Intersectional strata included education (’less than high school’ vs. ‘at least high school’), birth sex (’female’ vs. ‘male’), sexual orientation (’lesbian’, ‘gay’, ‘bisexual and others’), income level (’low’, ‘medium’, and ‘high’), and race/ethnicity (’Black’, ‘Hispanic’ and ‘White and others’). To address issues with sparse data, categories with small percentages (<5%) were combined with related groups to ensure robust statistical analysis.

### Analytical Procedures

*First,* descriptive analyses were conducted to summarize the data distributions of outcome variables (AUDIT and DUDIT) and explanatory variables. Frequencies and percentages were computed for categorical variables, while means and standard deviations were calculated for continuous variables. We further conducted stratified analyses to examine differ*ences across* key demographic subgroups (e.g., gender, sexual orientation, income level). Effect sizes (Cohen’s d and eta-squared) with 95% confidence intervals (CI) were calculated to assess the magnitude of group differences. *Secondly*, Bayesian hierarchical models were used to investigate associations between resilience and stressor predictors with substance use outcomes. Bayesian analysis was chosen for its flexibility in modeling complex hierarchical structures and its ability to incorporate prior information to improve model estimation, particularly when dealing with small or sparse data within intersectional strata. This approach allows for more robust estimates and credible intervals, accounting for uncertainty in parameter estimation.^18,36^ Random intercept models accounted for within-strata variability, while fixed effects estimated predictor-outcome relationships. Posterior distributions of coefficients were summarized using means, standard deviations, and 95% credible intervals. Convergence diagnostics (e.g., R-hat) and effective sample sizes (ESS) were evaluated to ensure model reliability. *Thirdly,* MAIHDA was conducted to assess heterogeneity in substance use outcomes across intersectional strata.

Null models estimated variance attributable to strata-level factors, and main-effects models incorporated demographic, resilience, and stressor predictors. Intraclass correlation coefficients (ICC) and proportional change in variance (PCV) quantified the explanatory power of the predictors. *Lastly*, multiple imputation was conducted using predictive mean matching via the mice package in R. Fifty imputations were performed to address missing data, and sparse strata (fewer than 10 participants) were merged using a lumping function to ensure sufficient group sizes for robust analysis. Imputation diagnostics confirmed the quality of the imputed datasets. All analyses were conducted using the ®R package.

## RESULTS

The descriptive analysis revealed notable differences in AUDIT and DUDIT scores across demographic groups. Females reported lower mean AUDIT scores (mean=2.61, standard deviation (sd) = 2.15) than males (mean=2.98, sd = 2.40; Cohen’s d = –0.16). Similarly, bisexual participants had higher mean DUDIT scores (mean=3.87, sd = 5.69) compared to lesbian participants (mean=2.30, sd = 4.70; eta-squared = 0.02). Resilience factors such as social support and life satisfaction exhibited modest variations across gender and sexual orientation. At the same time, stressors like everyday discrimination were consistently higher among racial/ethnic minorities and low-income groups (**Table 1**). When assessing strata-specific data and excluding outliers, the group labeled as Black bisexual male with medium income and at least high school education reported the highest AUDIT score (mean=7.00, sd=00), and Hispanic bisexual male with medium income and less than high school reported the highest DUDIT score (mean=25.00, sd=0.00) (**SuppTable 1a-1b**).

**Table 1.** Descriptive Analysis for substance use, resilience factors and stressors by demographic factors.

| Demographics |  | Substance use |  | Resilience Factors |  |  |  | Stressors |  |  |  |
| --- | --- | --- | --- | --- | --- | --- | --- | --- | --- | --- | --- |
|  |  | ADUIT | DUDIT | Connected<br>ness | Social well-being | Life<br>Satisfaction | Social<br>support | Everyday<br>discrimination | Felt stigma | Healthcare<br>Threat | Internalized<br>stigma |
| **Gender** | Female (n=805) | 2.61<br>(sd=2.15) | 3.30<br>(sd=5.56) | 2.97<br>(sd=0.56) | 4.58<br>(sd=0.92) | 4.31<br>(sd=1.65) | 5.25<br>(sd=1.27) | 1.99<br>(sd=0.71) | 2.70<br>(sd=0.92) | 2.52<br>(sd=1.01) | 1.54<br>(sd=0.69) |
|  | Male (n=691) | 2.98<br>(sd=2.40) | 3.30<br>(sd=5.41) | 2.94<br>(sd=0.57) | 4.74<br>(sd=0.88) | 4.33<br>(sd=1.60) | 5.14<br>(sd=1.32) | 1.81<br>(sd=0.66) | 2.62<br>(sd=0.96) | 2.63<br>(sd=1.13) | 1.73<br>(sd=0.82) |
|  | Cohen's d and 95 CI | **-0.16**<br>**(-0.27, -0.06)** | 2.29e-04<br>(-0.10, 0.10) | 0.05<br>(-0.05, 0.15) | **-0.18**<br>**(-0.28, -0.08)** | -0.01<br>(-0.11, 0.09) | 0.08<br>(-0.02, 0.18) | **0.27**<br>**(0.16, 0.37)** | 0.08<br>(-0.02, 0.18) | -0.10<br>(-0.20, 0.00) | **-0.25**<br>**(-0.35, -0.15)** |
| **Sexual**<br>**Orientation** | Bisexual (n=490) | 2.81<br>(sd=2.32) | 3.87<br>(sd=5.69) | 2.90<br>(sd=0.521) | 4.48<br>(sd=0.906) | 4.05<br>(sd=1.58) | 5.16<br>(sd=1.22) | 2.05<br>(sd=0.71) | 2.69<br>(sd= 0.93) | 2.46<br>(sd=1.03) | 1.67<br>(sd=0.79) |
|  | Gay (n=533) | 2.89<br>(sd=2.29) | 2.90<br>(sd=5.24) | 2.98<br>(sd=0.57) | 4.81<br>(sd=0.88) | 4.46<br>(sd=1.63) | 5.18<br>(sd=1.39) | 1.76<br>(sd=0.66) | 2.62<br>(sd=0.96) | 2.64<br>(sd=1.13) | 1.68<br>(sd=0.79) |
|  | Lesbian (n=288) | 2.48<br>(sd=2.13) | 2.30<br>(sd=4.70) | 3.01<br>(sd=0.60) | 4.73<br>(sd=0.97) | 4.55<br>(sd=1.67) | 5.32<br>(sd=1.30) | 1.81<br>(sd=0.66) | 2.73<br>(sd=0.91) | 2.50<br>(sd=0.96) | 1.52<br>(sd=0.68) |
|  | Others (n=207) | 2.86<br>(sd=2.36) | 4.52<br>(sd=6.76) | 3.01<br>(sd= 0.58) | 4.58<br>(sd=0.80) | 4.25<br>(sd=1.58) | 5.13 (sd=1.24) | 2.06 (sd=0.70) | 2.59<br>(sd=0.96) | 2.78<br>(sd=1.09) | 1.53<br>(sd=0.68) |
|  | Eta square | 4.36e-03<br>[0.00, 1.00] | **0.02**<br>**[0.01, 1.00]** | 6.81e-03<br>[0.00, 1.00] | **0.02**<br>**[0.01, 1.00]** | **0.02**<br>**[0.01, 1.00]** | 2.50e-03 \|<br>[0.00, 1.00] | **0.04**<br>**[0.02, 1.00]** | 2.61e-03<br>[0.00, 1.00] | **0.01**<br>**[0.00, 1.00]** | 8.76e-03 \|<br>[0.00, 1.00] |
| **Race** | Black (n=182) | 2.59<br>(sd=2.28) | 3.78<br>(sd=6.49) | 3.03<br>sd=0.598 | 4.37<br>sd=0.875 | 3.75<br>sd=1.63 | 4.96<br>(sd=1.35) | 2.25<br>(sd=0.77) | 2.86<br>(sd=0.86) | 2.59<br>(sd=1.15) | 1.81<br>(sd=0.87) |
|  | Hispanic (n=270) | 2.73<br>(sd= 2.19) | 4.00<br>(sd=6.26) | 3.07<br>(sd=0.53) | 4.58<br>(sd=0.84) | 4.26<br>(sd=1.54) | 5.36<br>(sd=1.18) | 1.96<br>(sd=0.67) | 2.65<br>(sd=0.91) | 2.69<br>(sd=1.07) | 1.69<br>(sd=0.81) |
|  | White and others<br>(n=1044) | 2.83<br>(sd=2.29) | 3.00<br>(sd=5.01) | 2.92<br>(sd=0.56) | 4.72<br>(sd=0.92) | 4.44<br>(sd=1.63) | 5.20<br>(sd=1.31) | 1.83<br>(sd=0.66) | 2.63<br>(sd=0.96) | 2.54<br>(sd= 1.05) | 1.57<br>(sd=0.72) |
|  | Eta square | 1.25e-03<br>[0.00, 1.00] | 6.01e-03<br>[0.00, 1.00] | **0.01**<br>**[0.00, 1.00]** | **0.02**<br>**[0.01, 1.00]** | **0.02**<br>**[0.01, 1.00]** | 6.90e-03<br>[0.00, 1.00] | 0.04<br>[0.03, 1.00] | 6.55e-03<br>[0.00, 1.00] | 2.68e-03<br>[0.00, 1.00] | 0.01<br>[0.00, 1.00] |
| **Income** | High (n=227) | 2.92<br>(sd=2.20) | 2.02<br>(sd=4.62) | 2.92<br>(sd=0.55) | 5.04<br>(sd=0.80) | 5.26<br>(sd=1.39) | 5.39<br>(sd=1.22) | 1.63<br>(sd=0.60) | 2.42<br>(sd=0.93) | 2.37<br>(sd=1.03) | 1.58<br>(sd=0.76) |
|  | Medium (n=397) | 3.03<br>(sd=2.33) | 2.41<br>(sd=4.23) | 2.89<br>(sd=0.58) | 4.85<br>(sd=0.89) | 4.72<br>(sd=1.56) | 5.26<br>(sd=1.38) | 1.76<br>(sd=0.61) | 2.61<br>(sd=0.91) | 2.53<br>(sd=1.07) | 1.60<br>(sd=0.73) |
|  | Low (n=894) | 2.64<br>(sd=2.27) | 4.05<br>(sd=6.16) | 3.00<br>(sd=0.56) | 4.47<br>(sd=0.89) | 3.90<br>(sd=1.57) | 5.11<br>(sd=1.28) | 2.04<br>(sd=0.72) | 2.75<br>(sd=0.95) | 2.65<br>(sd=1.07) | 1.65<br>(sd=0.77) |
|  | Eta square | 5.86e-03<br>[0.00, 1.00] | **0.03**<br>**[0.01, 1.00]** | 8.17e-03 \|<br>[0.00, 1.00] | **0.06**<br>**[0.04, 1.00]** | **0.10**<br>**[0.08, 1.00]** | 6.53e-03 \|<br>[0.00, 1.00] | **0.06**<br>**[0.04, 1.00]** | **0.02**<br>**[0.01, 1.00]** | 8.45e-03 \|<br>[0.00, 1.00] | 1.74e-03<br>[0.00, 1.00] |
| **Education** | Less than high school<br>(n=309) | 2.43<br>(sd=2.31) | 4.01<br>(sd=5.99) | 3.00<br>(sd=0.60) | 4.29<br>(sd=0.91) | 3.93<br>(sd=1.53) | 5.02<br>(sd=1.28) | 2.11 (sd=0.76) | 2.80<br>(sd=0.92) | 2.68<br>(sd=1.05) | 1.71<br>(sd=0.70) |
|  | At least high school<br>(n=1209) | 2.87<br>(sd=2.27) | 3.14<br>(sd=5.44) | 2.95<br>(sd=0.55) | 4.75<br>(sd=0.88) | 4.42<br>(sd=1.64) | 5.24<br>(sd=1.30) | 1.85 (sd=0.67) | 2.62<br>(sd=0.95) | 2.55<br>(sd=1.07) | 1.60<br>(sd=0.77) |
|  | Cohen's d and 95 CI | **-0.19**<br>**(-0.32, -0.07)** | **0.16**<br>**(0.03, 0.28)** | 0.09<br>(-0.04, 0.21) | -0.51<br>(0.64, -0.38) | **-0.30**<br>**(-0.43, -0.17)** | **-0.17**<br>**(-0.29, -0.04)** | **0.37**<br>**(0.25, 0.50)** | **0.18 (0.06,**<br>**0.31)** | **0.12**<br>**(0.00, 0.25)** | **0.14**<br>**(0.02, 0.27)** |

Bayesian regression analyses highlighted the significant role of everyday discrimination as a predictor of both alcohol and drug use. For AUDIT, everyday discrimination had a positive association (Estimate = 0.10, 95% CI [0.01, 0.18]), while perceived stigma showed a negative trend (Estimate = –0.06, 95% CI [-0.12, 0.00]). For DUDIT, everyday discrimination exhibited a more substantial effect (Estimate = 0.23, 95% CI [0.15, 0.32]), with perceived stigma negatively associated (Estimate = –0.11, 95% CI [-0.17, –0.05]). Resilience factors such as social well-being demonstrated protective effects against drug use (Estimate = –0.17, 95% CI [-0.23, –0.10]) (**Table 2**). Speaking of the model fit, the observed data (y) closely align with the replicated distributions (y_rep), indicating that the models effectively captured the data’s underlying structure for both models (**SuppFig.1a-1b**). The posterior distributions of predictors revealed that everyday discrimination consistently demonstrated the strongest positive association with both outcomes. On the other hand, resilience factors such as social well-being showed protective effects, particularly against drug use in the DUDIT model. Negative associations were observed for perceived stigma, highlighting its complex and varying role across the models (**Fig.1a-1b**).

**Table 2.** Bayesian Regression Coefficients for Stressor and Resilience Factors Predicting Alcohol Use (AUDIT) and Drug Use (DUDIT) Among Sexual Minority Groups.

| Predictor | Estimate | Est. Error | 95% CI (Lower) | 95% CI (Upper) | Rhat | Bulk ESS | Tail ESS |
| --- | --- | --- | --- | --- | --- | --- | --- |
| **ALCOHOL USE AS THE OUTCOME (AUDIT)** |  |  |  |  |  |  |  |
| Connectedness | 0.02 | 0.05 | -0.08 | 0.11 | 1.0 | 11209 | 6138 |
| Social Well-Being | 0.02 | 0.04 | -0.05 | 0.08 | 1.0 | 10301 | 6961 |
| Life Satisfaction | 0.03 | 0.02 | -0.01 | 0.07 | 1.0 | 10304 | 6517 |
| Social Support | -0.0 | 0.02 | -0.05 | 0.04 | 1.0 | 10639 | 6198 |
| Everyday Discrimination | 0.10 | 0.04 | 0.01 | 0.18 | 1.0 | 10111 | 6240 |
| Perceived Stigma | -0.06 | 0.03 | -0.12 | 0.0 | 1.0 | 10760 | 5672 |
| Healthcare Threat | 0.01 | 0.03 | -0.05 | 0.06 | 1.0 | 11481 | 5983 |
| Internalized Stigma | -0.04 | 0.04 | -0.11 | 0.04 | 1.0 | 11831 | 6024 |
| Connectedness | 0.09 | 0.05 | -0.0 | 0.18 | 1.0 | 10742 | 6382 |
| Social Well-Being | -0.17 | 0.04 | -0.23 | -0.10 | 1.0 | 9660 | 5296 |
| Life Satisfaction | -0.01 | 0.02 | -0.05 | 0.03 | 1.0 | 7872 | 5683 |
| Social Support | 0.01 | 0.02 | -0.03 | 0.05 | 1.0 | 10568 | 5696 |
| Everyday Discrimination | 0.23 | 0.04 | 0.15 | 0.32 | 1.0 | 9700 | 6046 |
| Perceived Stigma | -0.11 | 0.03 | -0.17 | -0.05 | 1.0 | 11429 | 5953 |
| Healthcare Threat | -0.05 | 0.03 | -0.10 | -0.00 | 1.0 | 11437 | 6222 |
| Internalized Stigma | 0.03 | 0.04 | -0.03 | 0.10 | 1.0 | 12347 | 5923 |

**Notes:**
- **Estimate:** Posterior mean of the regression coefficient, indicating the central tendency of the posterior distribution. - **Est. Error:** Standard deviation of the posterior distribution, reflecting uncertainty around the estimate. - **Credible Interval (l-95% CI, u-95% CI):** 95% credible interval showing the range within which the parameter likely falls with 95% probability. Indicates statistical or practical relevance (whether it includes 0). - **Rhat:** Convergence diagnostic; values near 1 indicate good convergence. - **Bulk ESS & Tail ESS:** Effective sample sizes assessing posterior sample quality. *Bulk ESS* reflects mixing in the bulk, while *Tail ESS* assesses reliability in the tails.

The MAIHDA analyses provided critical insights into substance use disparities by examining the contributions of demographic factors (**Table 3a**), and resilience factors and stressors (**Table 3b**). In Table 3a, models incorporating demographic factors such as birth sex, sexual orientation, and race reduced the unexplained strata-level variation in AUDIT to 5.89% and DUDIT to 0%. Birth sex and sexual orientation emerged as significant explanatory variables, with male and bisexual participants demonstrating higher substance use outcomes compared to other groups. Table 3b highlights the role of resilience factors and stressors in explaining substance use outcomes. Everyday discrimination was the strongest positive predictor of AUDIT (Estimate = 0.097, 95% CI [0.013, 0.182]) and DUDIT (Estimate = 0.233, 95% CI [0.151, 0.315]), while social well-being exhibited significant protective effects against drug use (Estimate = –0.166, 95% CI [-0.234, –0.097]). Perceived stigma also had a consistent negative association with both outcomes. These key explanatory factors explained 93.6% of the strata-level variation in AUDIT and 34.45% in DUDIT, underscoring their substantial contributions to understanding substance use disparities across intersectional strata. Our analyses revealed that main-effects models with predictors (e.g., everyday discrimination) significantly reduce strata-level variation compared to null models, especially for DUDIT outcomes (**Table 3a-3b**, **Fig.2**).

**Table 3a.** Multilevel Analysis of Individual Heterogeneity and Discriminatory Accuracy (MAIHDA) with Demographic Predictors.

|  | **AUDIT** |  | **DUDIT** |  |
| --- | --- | --- | --- | --- |
|  | Null | Main-effect model | Null | Main-effect model |
| **Fixed effect (estimate and 95% CI)** |  |  |  |  |
| *Intercept* | -0.009 (-0.074, 0.055) | -0.150 (-0.402, 0.103) | 0.027 (-0.053, 0.108) | -0.125 (-0.371, 0.121) |
| **Gender** |  |  |  |  |
| Female | -- |  | -- |  |
| Male | -- | 0.245 (0.081, 0.409) | -- | 0.070 (-0.090, 0.229) |
| **Sexual Orientation** |  |  |  |  |
| Bisexual | -- | ref | -- | ref |
| Gay | -- | -0.202 (-0.373, -0.031) | -- | -0.127 (-0.294, 0.041) |
| Lesbian | -- | -0.109 (-0.268, 0.050) | -- | -0.179 (-0.333, -0.025) |
| Others | -- | -0.005 (-0.174, 0.164) | -- | 0.102 (-0.063, 0.266) |
| **Race** |  |  |  |  |
| Black | -- | ref | -- |  |
| Hispanic | -- | -0.011 (-0.201, 0.180) | -- | 0.025 (-0.162, 0.212) |
| White and others | -- | 0.028 (-0.133, 0.188) | -- | -0.125 (-0.283, 0.032) |
| **Income** |  |  |  |  |
| High | -- | ref | -- |  |
| Medium | -- | 0.049 (-0.120, 0.219) | -- | 0.111 (-0.054, 0.275) |
| Low | -- | -0.096 (-0.255, 0.063) | -- | 0.359 (0.205, 0.514) |
| **Education** |  |  |  |  |
| Less than high school | -- | ref | -- |  |
| At least high school | -- | 0.195 (0.063, 0.327) | -- | -0.004 (-0.133, 0.125) |
| **Random Effect (estimate and 95% CI)** |  |  |  |  |
| *Strata* | 0.016 (-0.107, 0.139) | 0.001 (-0.029, 0.031) | 0.0477 (-0.166, 0.261) | n/a |
| *Residual* | 0.987 (0.014, 1.961) | 0.985 (0.012, 1.958) | 0.958 (-0.001, 1.917) | 0.963 (0.001, 1.925) |
| Percent of Between-Strata Variation Unexplained by Main Effects | n/a | 5.89% | n/a | 0% |
| ICC | 1.72 % | 0.1 % | 4.93 % | 0.09 % |
| PCV |  | 94.16 % |  | 98.24 % |

Notes: ICC: intra-cluster correlation; PCV: proportional change in variance

**Table 3b.**
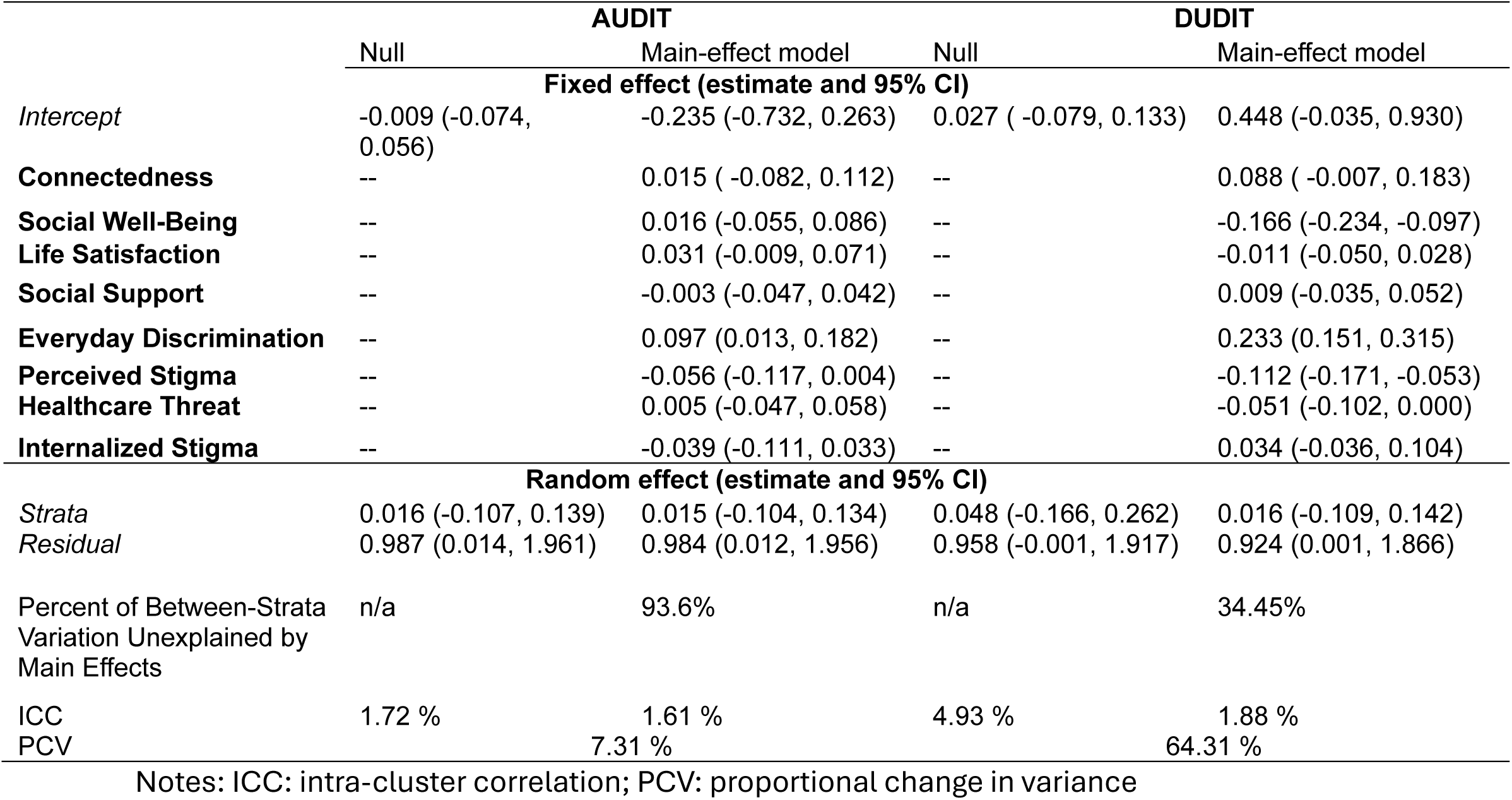
Multilevel Analysis of Individual Heterogeneity and Discriminatory Accuracy (MAIHDA) with Resilience and Stressor Predictors.

## DISCUSSION

Our study employed the MAIHDA to assess the nuanced associations between resilience factors, stressors, and alcohol and drug use behaviors among SMPs, guided by the intersectional framework.^12,14^ We quantified how combinations of social identities modulate these associations to explore the interplay of resilience and stress across diverse intersectional strata. The analyses revealed distinct patterns in how resilience and stressor variables relate to alcohol and drug use outcomes among SMPs. Social support and community connectedness consistently demonstrated protective effects against substance use across strata, while the magnitude of these associations varied. Stressors such as everyday discrimination and internalized homophobia were strongly associated with increased substance use, highlighting the role of minority stress in driving health disparities. Notably, our MAIHDA framework identified significant heterogeneity in these associations across intersectional strata, emphasizing the critical influence of individual identities such as sex, race/ethnicity, sexual orientation, income level, and education.

Our study echoed findings from existing studies that reported social support and community connectedness played a buffering role in mitigating substance use among socially marginalized groups.^30,37^ Furthermore, stressors such as everyday discrimination and internalized homophobia showed strong associations with increased substance use, consistent with the minority stress framework, which posits that chronic stress from social marginalization exacerbates health disparities.^32^ Notably, the strength of these associations was intensified in intersectional strata characterized by compounding vulnerabilities, such as low-income, non-White, and gender-minority groups. Similar patterns were reported in studies conducted by Coulter et al. (2018) and Mereish & Bradford (2014), where intersectional stressors disproportionately impacted substance use among SMPs.^38,39^ Our findings extend the existing literature by leveraging the MAIHDA framework to capture heterogeneity in these associations across intersectional strata.

Unlike traditional models, which often assume uniform stress and resilience effects, the MAIHDA application captured previously unrecognized variability in the protective effects of resilience factors, underscoring the necessity of intersectional analyses. Specifically, while resilience factors such as social well-being offered robust protection against substance use for higher-income, White SMPs, their protective effects were less pronounced in low-income, non-White groups. This observation underscores the necessity of addressing structural determinants that shape the efficacy of resilience factors. Similarly, the amplified effects of stressors in economically marginalized and racially ethnically diverse strata highlight the intersectionalized vulnerabilities faced by SMPs at the intersection of multiple minority identities. Therefore, MAIHDA identified substantial between-strata heterogeneity, with intersectional strata explaining a meaningful proportion of variance in substance use outcomes.

Several limitations warrant consideration. *First*, the cross-sectional design precludes causal inference, limiting our ability to disentangle temporal relationships between stressors, resilience factors, and substance use. *Second*, the reliance on self-reported measures may introduce recall bias. *Third*, while MAIHDA allows for a detailed examination of intersectionality, some strata had sparse data, necessitating strata merging that may obscure finer distinctions. *Fourth*, some strata suffer sparse data issues due to the limited sample size. *Fifth*, the nature of secondary data analyses may constrain the ability to test hypotheses beyond the scope of the original data collection, as the available variables may not fully capture all relevant variables or account for confounders critical to the research question. *Lastly*, the study focused on SMPs in the U.S., and findings may not generalize to other cultural or geographic contexts.

Despite these limitations, our findings underscore the importance of incorporating intersectional analyses into health research with SMPs. Our study highlights the necessity of tailored interventions that address the unique needs of SMP subgroups, particularly those facing intersecting structural and social disadvantages. For example, interventions should prioritize reducing stressors like everyday discrimination while strengthening resilience through community support initiatives. Policymakers should also consider tackling structural inequities, such as income disparities and systemic racism, that exacerbate substance use risks among SMPs. Future research should build on this study by employing longitudinal designs to assess causal pathways and changes over time. Additionally, integrating qualitative methods could deepen our understanding of lived experiences within specific strata. Expanding MAIHDA to examine other health outcomes and incorporating more diverse SMP populations would further enhance the generalizability and impact of this work.

In conclusion, this study demonstrates the utility of MAIHDA for examining health disparities through an intersectional lens among SMPs. By capturing the complex interplay of resilience, stress, and social identities, our findings provide actionable insights for advancing health equity among SMPs and addressing the disproportionate burden of substance use in this vulnerable population.

## Data Availability

Data are extracted from the public health repository ICPSR website. All data are deidentified and are publicly accessible.

https://www.icpsr.umich.edu/web/DSDR/studies/37166

## Authors’ Statement

Zhang and Liu are leading the data analyses and development of the manuscript, Qiao and Chang contributed to conceptual development, manscript refinement and interpretation of findings. All authors reviewed and approved the final manuscript and contributed to shaping the study’s direction and ensuring its methodological rigor.

## Data statement

All data were downloaded from the data repository from https://www.icpsr.umich.edu/web/DSDR/studies/37166

## Aritcle Summary

### Strengths and Limitations

- This study employs Multilevel Analysis of Individual Heterogeneity and Discriminatory Accuracy (MAIHDA), a robust statistical method that captures the nuanced interplay of intersecting identities, allowing for a detailed understanding of resilience and stressor effects across diverse sexual minority populations (SMPs).
- The study leverages validated instruments (e.g., AUDIT, DUDIT) for substance misuse and well-established scales for resilience and stressors, ensuring reliability and consistency in measuring key variables.
- By using data from the Generation Study (https://www.generations-study.com/), which includes a diverse, nationally representative cohort, the findings are broadly applicable to SMPs in the United States.
- The study’s cross-sectional nature limits causal inferences, restricting the ability to establish temporal relationships between stressors, resilience, and substance misuse.
- While MAIHDA addresses sparse data issues to an extent, merging small intersectional strata may obscure finer distinctions in resilience and stressor impacts, potentially limiting the granularity of the findings.

### The original protocol for the study

see the supplementary files and the study website (https://www.generations-study.com/).

### Ethical Approval

Waived as it is secondary data analysis

### Informed consent

Not Applicable

### Patient and Public Involvement statement

It is a secondary data analysis using data that are publicly available. Therefore, it is not appropriate or possible to involve patients or the public in the design, or conduct, or reporting, or dissemination plans of our research.

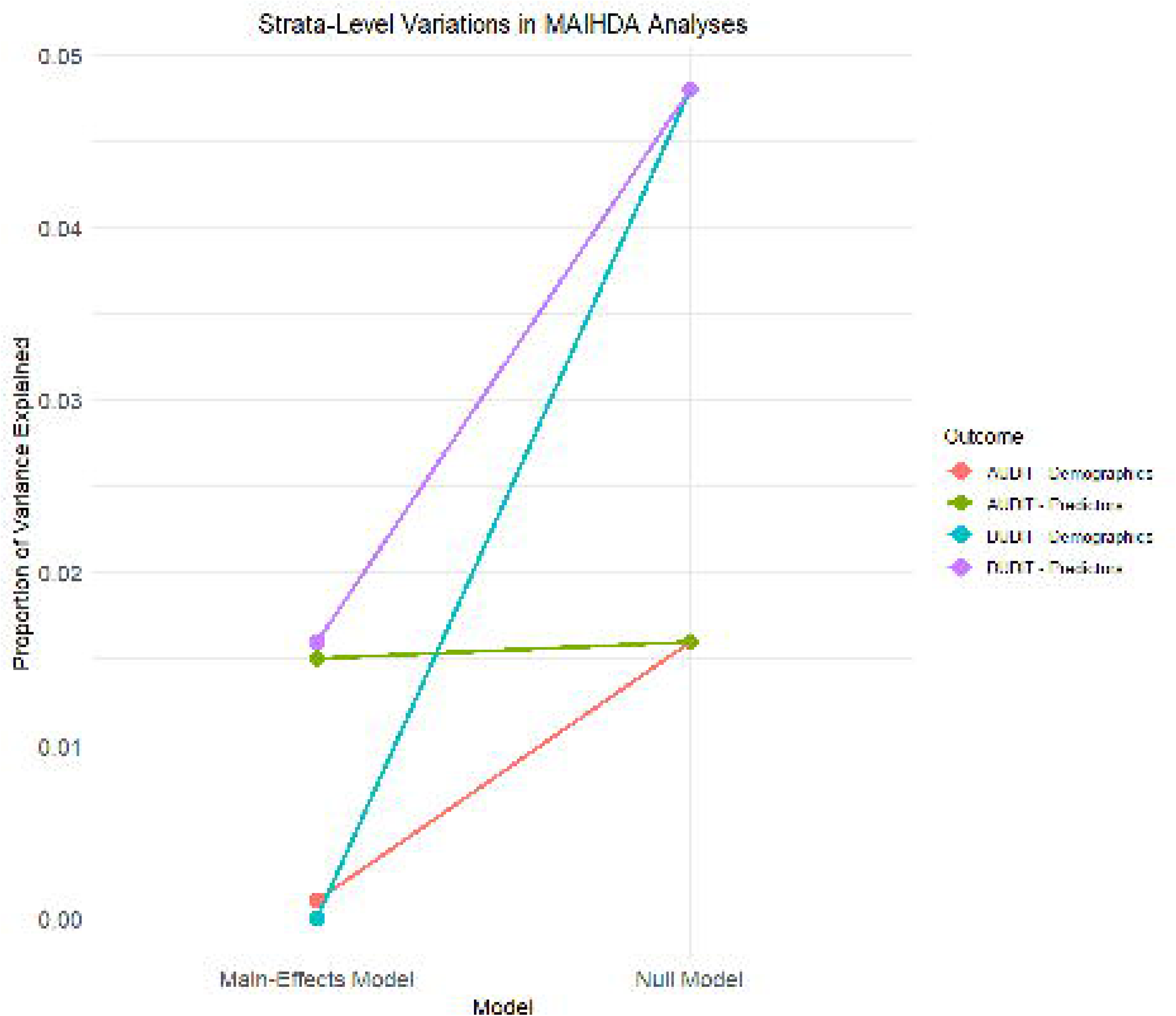

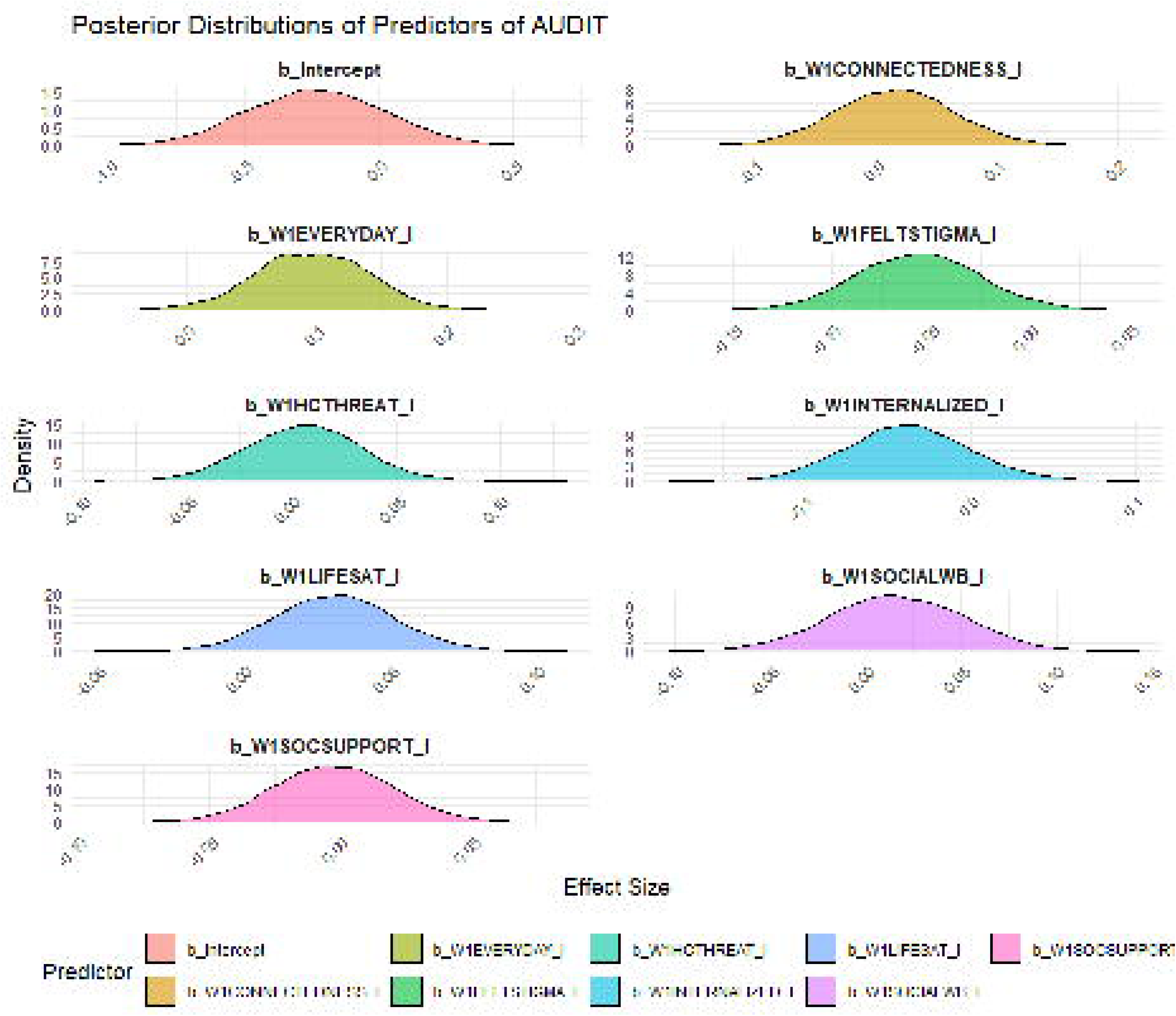

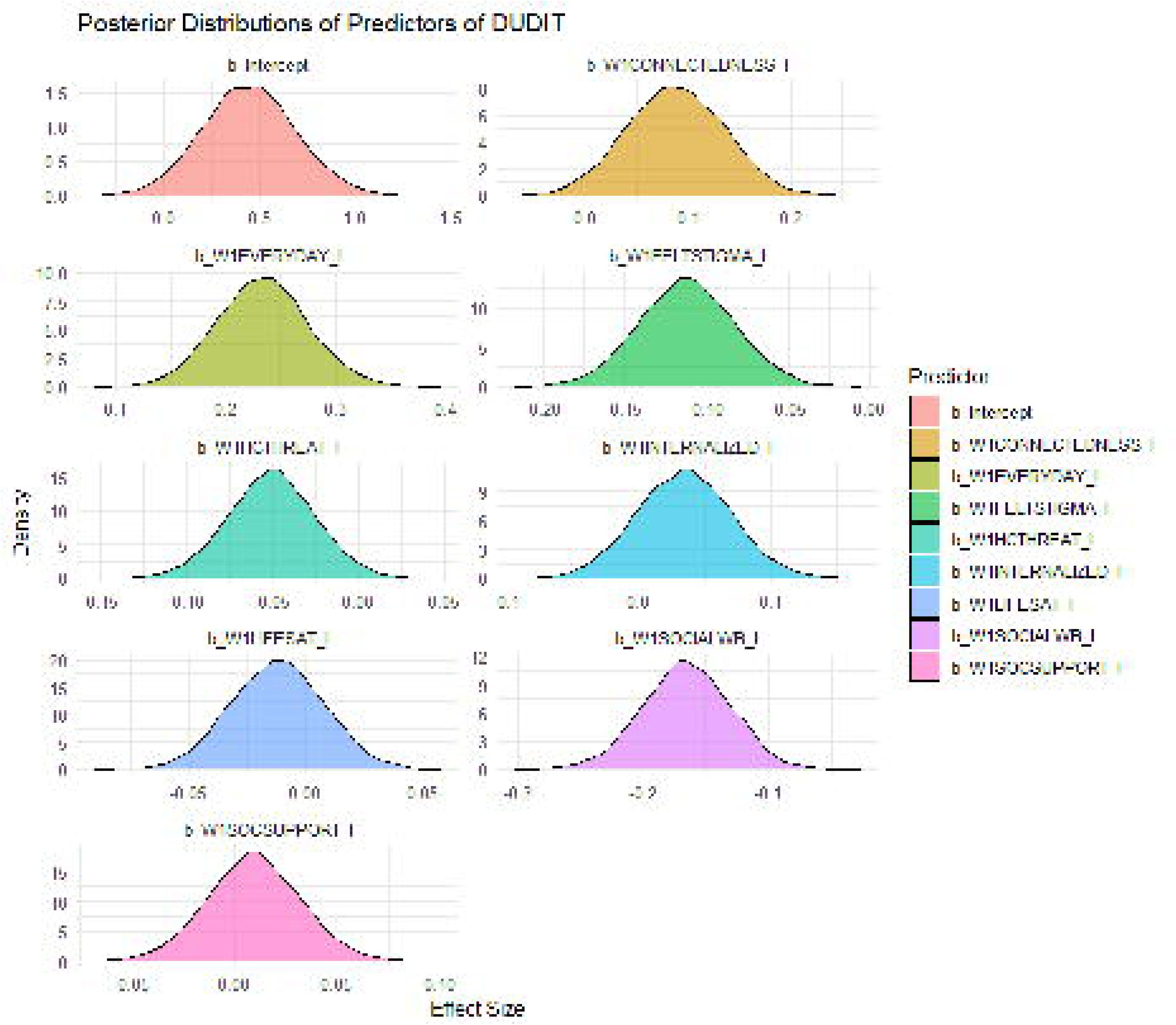

## REFERENCES

1. SAMHSA. Results from the 2013 National Survey on Drug Use and Health: Summary of National Findings: DHHS; 2014.

2. SAMHSA. Center for Behavioral Health Statistics and Quality, National Survey on Drug Use and Health, 2022. 2023; https://www.samhsa.gov/data/data-we-collect/nsduh-national-survey-drug-use-and-health.

3. Schuler MS, Prince DM, Breslau J, Collins RL. Substance Use Disparities at the Intersection of Sexual Identity and Race/Ethnicity: Results from the 2015-2018 National Survey on Drug Use and Health. LGBT health. Aug/Sep 2020;7(6):283–291.

4. Meyer IH. Minority stress and mental health in gay men. Journal of health and social behavior. Mar 1995;36(1):38–56.

5. Brooks BD, Kaniuka A, Job SA, et al. Anticipated Sexual Minority Stress and Mental Health after the 2016 Presidential Election: Examining a Psychological Mediation Framework. Journal of homosexuality. Nov 10 2023;70(13):3125–3148.

6. Katz-Wise SL, Sarda V, Austin SB, Harris SK. Longitudinal effects of gender minority stressors on substance use and related risk and protective factors among gender minority adolescents. Plos one. 2021;16(6):e0250500.

7. Ungar M. Resilience and culture: The diversity of protective processes and positive adaptation. Youth resilience and culture: Commonalities and complexities. 2015:37–48.

8. Harper GW, Bruce D, Hosek SG, Fernandez MI, Rood BA, Interventions AMTNfHA. Resilience processes demonstrated by young gay and bisexual men living with HIV: Implications for intervention. AIDS patient care and STDs. 2014;28(12):666–676.

9. Motley DN, Victorian J, Denis K, Brooks BD. Applying an intersectionality framework to health services research. Families, systems & health: the journal of collaborative family healthcare. Dec 2023;41(4):417–424.

10. Green MA, Evans CR, Subramanian SV. Can intersectionality theory enrich population health research? Social science & medicine (1982). Apr 2017;178:214–216.

11. Turan JM, Elafros MA, Logie CH, et al. Challenges and opportunities in examining and addressing intersectional stigma and health. BMC medicine. Feb 15 2019;17(1):7.

12. Ghasemi E, Majdzadeh R, Rajabi F, et al. “Applying Intersectionality in designing and implementing health interventions: a scoping review”. BMC public health. Jul 16 2021;21(1):1407.

13. Evans CR. Adding interactions to models of intersectional health inequalities: Comparing multilevel and conventional methods. Social science & medicine (1982). Jan 2019;221:95–105.

14. Alvarez CH, Evans CR. Intersectional environmental justice and population health inequalities: A novel approach. Social science & medicine (1982). Jan 2021;269:113559.

15. Evans CR. Modeling the intersectionality of processes in the social production of health inequalities. Social science & medicine (1982). Apr 2019;226:249–253.

16. Evans CR, Williams DR, Onnela JP, Subramanian SV. A multilevel approach to modeling health inequalities at the intersection of multiple social identities. Social science & medicine (1982). Apr 2018;203:64–73.

17. Merlo J. Invited commentary: multilevel analysis of individual heterogeneity-a fundamental critique of the current probabilistic risk factor epidemiology. Am J Epidemiol. Jul 15 2014;180(2):208–212; discussion 213-204.

18. Merlo J. Multilevel analysis of individual heterogeneity and discriminatory accuracy (MAIHDA) within an intersectional framework. Social science & medicine (1982). Apr 2018;203:74–80.

19. Khalaf K, Axelsson Fisk S, Ekberg-Jansson A, Leckie G, Perez-Vicente R, Merlo J. Geographical and sociodemographic differences in discontinuation of medication for Chronic Obstructive Pulmonary Disease – A Cross-Classified Multilevel Analysis of Individual Heterogeneity and Discriminatory Accuracy (MAIHDA). Clinical epidemiology. 2020;12:783–796.

20. Persmark A, Wemrell M, Zettermark S, Leckie G, Subramanian SV, Merlo J. Precision public health: Mapping socioeconomic disparities in opioid dispensations at Swedish pharmacies by Multilevel Analysis of Individual Heterogeneity and Discriminatory Accuracy (MAIHDA). PloS one. 2019;14(8):e0220322.

21. Meyer IH. Generations: A Study of the Life and Health of LGB People in a Changing Society, United States, 2016-2019: Inter-university Consortium for Political and Social Research [distributor]; 2023.

22. Bush K, Kivlahan DR, McDonell MB, Fihn SD, Bradley KA. The AUDIT alcohol consumption questions (AUDIT-C): an effective brief screening test for problem drinking. Ambulatory Care Quality Improvement Project (ACQUIP). Alcohol Use Disorders Identification Test. Archives of internal medicine. Sep 14 1998;158(16):1789–1795.

23. Higgins-Biddle JC, Babor TF. A review of the Alcohol Use Disorders Identification Test (AUDIT), AUDIT-C, and USAUDIT for screening in the United States: Past issues and future directions. The American journal of drug and alcohol abuse. 2018;44(6):578–586.

24. Reinert DF, Allen JP. The Alcohol Use Disorders Identification Test (AUDIT): a review of recent research. Alcoholism, clinical and experimental research. Feb 2002;26(2):272–279.

25. Hildebrand M. The Psychometric Properties of the Drug Use Disorders Identification Test (DUDIT): A Review of Recent Research. Journal of substance abuse treatment. Jun 2015;53:52–59.

26. Neufeld J, Ullrich F, Merchant KAS, et al. Change in Drug Use Disorders Identification Test – Consumption (DUDIT-C) with Telehealth Treatment Compared to in-Person Treatment. Substance use & misuse. 2023;58(9):1168–1171.

27. Zimet GD, Powell SS, Farley GK, Werkman S, Berkoff KA. Psychometric characteristics of the Multidimensional Scale of Perceived Social Support. Journal of personality assessment. Winter 1990;55(3-4):610–617.

28. Diener E, Emmons RA, Larsen RJ, Griffin S. The Satisfaction With Life Scale. Journal of personality assessment. Feb 1985;49(1):71–75.

29. Keyes CLM. Social Well-Being. Social Psychology Quarterly. 1998;61:121–140.

30. Frost DM, Meyer IH. Measuring community connectedness among diverse sexual minority populations. Journal of sex research. 2012;49(1):36–49.

31. Williams DR, Yan Y, Jackson JS, Anderson NB. Racial Differences in Physical and Mental Health: Socioeconomic Status, Stress and Discrimination. Journal of health psychology. Jul 1997;2(3):335–351.

32. Meyer IH. Prejudice, social stress, and mental health in lesbian, gay, and bisexual populations: conceptual issues and research evidence. Psychological bulletin. Sep 2003;129(5):674–697.

33. Herek GM. Sexual stigma and sexual prejudice in the United States: a conceptual framework. *Nebraska Symposium on Motivation*. Nebraska Symposium on Motivation. 2009;54:65–111.

34. Fingerhut A, Abdou CM. The Role of Healthcare Stereotype Threat and Social Identity Threat in LGB Health Disparities. Journal of Social Issues. 2017;73(3):493–507.

35. Theodore JL, Shidlo A, Zemon V, et al. Psychometrics of an internalized homophobia instrument for men. Journal of homosexuality. 2013;60(4):558–574.

36. Kruschke JK. Doing Bayesian Data Analysis A Tutorial with R, JAGS, and Stan. London, UK: Elsevier 2015.

37. Hatzenbuehler ML, Phelan JC, Link BG. Stigma as a fundamental cause of population health inequalities. American journal of public health. May 2013;103(5):813–821.

38. Coulter RW, Birkett M, Corliss HL, Hatzenbuehler ML, Mustanski B, Stall RD. Associations between LGBTQ-affirmative school climate and adolescent drinking behaviors. Drug and alcohol dependence. Apr 1 2016;161:340–347.

39. Mereish EH, Bradford JB. Intersecting identities and substance use problems: sexual orientation, gender, race, and lifetime substance use problems. Journal of studies on alcohol and drugs. Jan 2014;75(1):179–188.

